# Effectiveness of an Ozone Disinfecting and Sanitizing Cabinet to Decontaminate a Surrogate Virus for SARS-CoV-2 on N-95 Masks

**DOI:** 10.1101/2020.11.04.20226233

**Authors:** Megan S. Beaudry, Julia C. Frederick, Megan E. J. Lott, William A. Norfolk, Travis C. Glenn, Erin K. Lipp

## Abstract

Medical demands during the COVID-19 pandemic have triggered a grave shortage of medical-grade personal protective equipment (PPE), especially, N95 respirators. N95 respirators are critical for the personal protection of medical providers and others when being exposed to individuals with infections caused by the SARS-CoV-2 coronavirus. To address the shortage of N95 respirators, innovative methods are needed to decontaminate coronaviruses from N95 respirators, allowing them to be safely reused by healthcare workers. For this research, we use a commercial ozone disinfecting cabinet to examine the efficacy of ozone-based disinfection of a conservative surrogate virus for SARS-CoV-2, the MS2 bacteriophage. Treatment of mask materials with enhanced ozone treatment resulted in 2.38-log _10_ (>99%) reduction of phage from household dust masks and a range of 1.43-log _10_ (96.2%) to 4-log _10_ (99.99%) reductions of phage from common N95 mask materials.

## Introduction

The COVID-19 pandemic, caused by the virus SARS-CoV-2, is a threat to public health worldwide. SARS-CoV-2 is an enveloped positive-sense, single-stranded RNA virus that first emerged in the Chinese province of Hubei in December 2019 [1]. SARS-CoV-2 causes a respiratory infection which is primarily transmitted via droplets and aerosols [1]. As of October 2020, COVID-19 deaths exceed 200,000 in the US and 1,000,000 globally [2].

The COVID-19 pandemic has substantially reduced the supply of medical grade PPE, specifically for filtering facepiece respirators (FFRs) (e.g., N95 masks). N95 masks are commonly used in infection control, as they are certified to filter 95% of airborne particles down to 0.3 microns. The World Health Organization (WHO) estimated that for each month the pandemic persists, 89 million medical-grade masks will be needed to respond to the outbreak [3]. Further compounding this problem, is the increased public demand for FFRs, depleting the stocks available to hospitals and other essential services. There is a substantial need for safe and effective methods of decontamination to ensure the supply of PPE can be maintained until resolution of the pandemic.

Throughout the pandemic, a number of academic institutions have collaborated with industry partners to find effective methods to decontaminate medical grade PPE with varying degrees of success [4, 5]. Here, we have used a commercial ozone disinfection cabinet to test the effect of two different ozone treatments on the survival of the MS2 bacteriophage. A non-enveloped, single-stranded RNA virus, the MS2 bacteriophage has been proposed as a conservative surrogate for the SARS-CoV-2 virus, as non-enveloped viruses are more resistant to decontamination than enveloped, lipid viruses [5, 6].

## Materials and Methods

### Propagation and Quantification of MS2 Phage

MS2 bacteriophage was kindly provided by Dr. Mark Sobsey (University of North Carolina – Chapel Hill) and was propagated according to methods described in the USEPA Manual of Methods for Virology. Briefly, viruses were propagated by inoculating 1 mL of rehydrated phage into a log-phase culture of the bacterial host, *Escherichia coli* strain C-3000 (ATCC® 15597), in tryptic soy broth and incubating for 6 h at 37°C with shaking. The culture was filtered through a 0.22 uM filter and the lysate coliphage stock was stored at 4°C. The concentration of the coliphage stock was quantified by the double-agar method. In this method, a tube of molten (0.7% TSA) “top agar” with 0.1 mL of log-phase host bacteria (*E. coli* C-3000) is inoculated with 1 mL of sample and poured onto a 1.5% TSA “bottom agar” plate. Plates were inverted and incubated at 37°C overnight.

### Mask deconstruction

N95 masks (3M™ Particulate Respirator 8511) were cut into 1 cm^2^ coupons (pieces) and divided into material groups: N95 mask filter material (i.e., the main filtration materials), N95 exhalation valve material (i.e., the filter within N95 exhalation valves), and the N95 elastic band. Additionally, a dust mask (3M™ Home Dust Mask) was cut into 1 cm^2^ coupons to make the test material. Only the main filtration material of this mask was tested. The layers of the main filtration materials of the masks were kept together.

### Coupon Inoculation

The MS2 coliphage stock was diluted in 1.5% beef extract (pH 7.5) to 10^5^ PFU/10 uL. An aliquot of 10 uL of the diluted phage stock was inoculated onto each coupon. For the main filtration materials of the masks (N95 and Dust Mask) the outermost layer was inoculated. The inoculum was set to dry at room temperature in a biosafety cabinet for 15 minutes.

### Coupon Treatment

A commercially available ozone disinfection cabinet was used to treat coupon materials following MS2 inoculation. Coupons were untreated, treated in the cabinet on the standard cycle, or treated in the cabinet on the enhanced cycle. The standard cycle consisted of 5 minutes and 49 seconds of ramp up where ozone concentration went from 149-19918 ppm linear, followed by 16 minutes and 26 seconds of peak hold at 20069 ppm linear, and 11 minutes and 47 seconds of ramp down from 18291 to 336 ppm linear. For the enhanced cycle, the ramp up period lasted for 14 minutes and 18 seconds where the ozone concentration went from 1906-35721 ppm linear, followed by 43 minutes and 53 seconds of peak hold at 39528 ppm linear, and 12 minutes and 38 seconds of ramp down time where ozone concentrations went from 37768 – 434 ppm linear.

### Coupon Elution

Using sterile forceps, each coupon was placed into sterile screw-cap tubes containing 4 mL of 1.5% Beef Extract. Phage was eluted from coupons by shaking on a platform for 20 min at 125 rpm (New Brunswick Scientific C24 Shaker Incubator), at room temperature. The eluent was diluted in series in 1.5% Beef Extract. Phage was quantified from the eluent using the double-agar method as described previously. At least three dilutions of each sample eluent were analyzed in triplicate to yield plaque counts of 30 to 80 PFU. Plates were inverted and incubated at 37°C overnight.

### Data analysis

Recovered PFUs were quantified by direct counts and analyzed using R statistical software [7]. Log reductions were calculated as the difference between the recovered PFUs from control and treated materials. Shapiro-Wilk tests were used to determine data normality. T-tests (α = 0.01) were used to compare significant differences between the log recovered PFUs from control coupon and treated coupons.

## Results

We used a disinfecting cabinet to test the effect of two different ozone treatments (standard treatment and enhanced treatment with double the exposure time and dose of O^3^) on the survival of a non-enveloped single-stranded RNA bacteriophage virus (MS2). The MS2 bacteriophage was inoculated on four materials: one from a common house dust mask and three materials commonly used in the construction of N95 masks (main filter material, elastic, and exhalation valve). The results show a substantial decrease in the level of recovered PFUs from treated mask materials. The dust mask shows a reduction of 2.50 and 2.38 for the standard and enhanced treatment, respectively. The N95 filter media shows a log reduction of 0.75 for the standard treatment, and 3.38 for the enhanced treatment. For the N95 exhalation valve we had a log reduction of 1.05 for the standard and 1.43 enhanced treatments. Lastly, for the N95 elastic there was a 3.95 and 4.00 log reduction for the standard and enhanced treatments, respectively. Log recovered PFUs were normally distributed for all samples according to the Shapiro-Wilk test. Viral recovery was statistically significant (p < 0.01) for all materials treated on the enhanced setting as well as standard treatments for the dust mask and N95 elastic.

**Figure 1:**
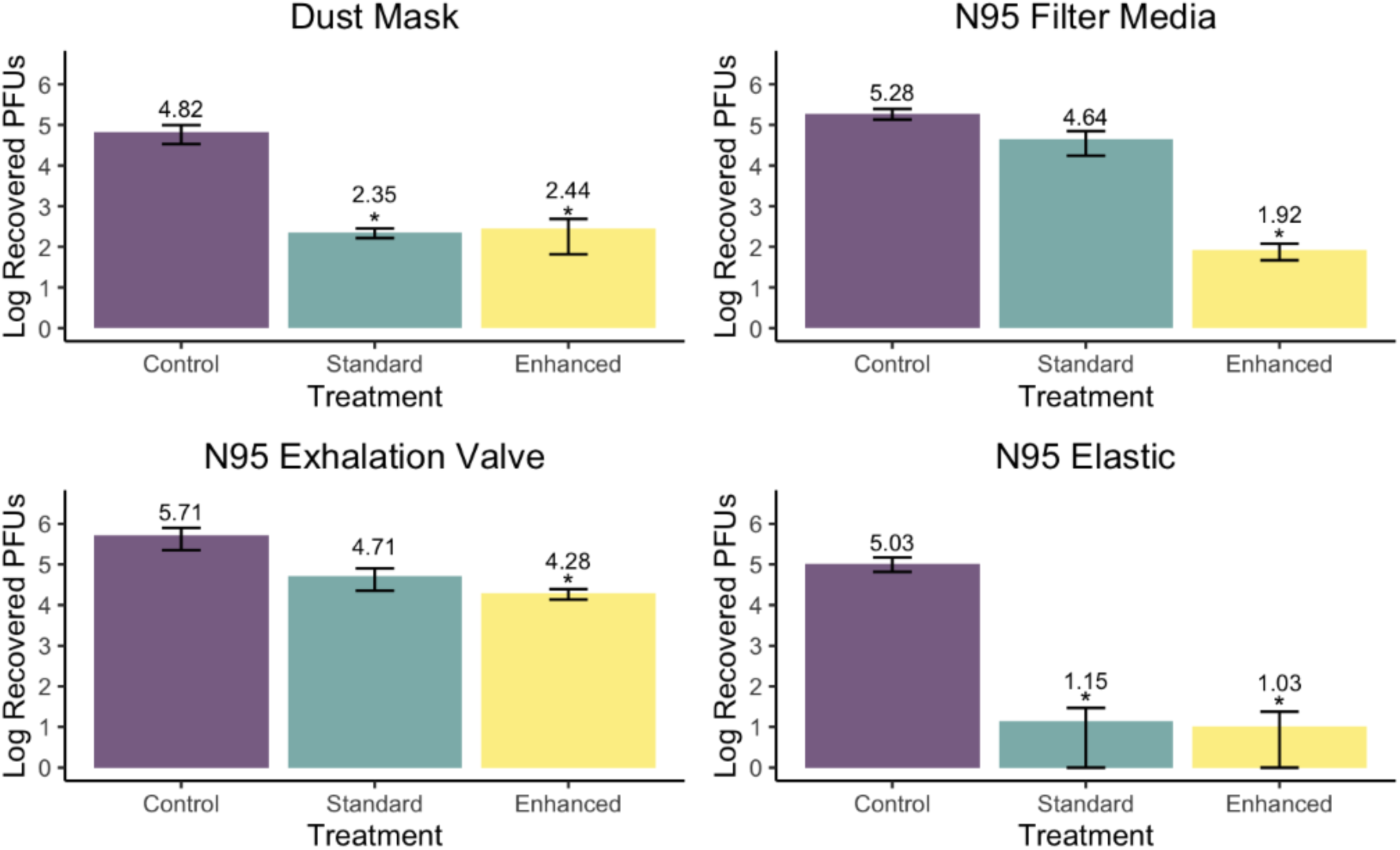
MS2 viruses enumerated from coupons. Three coupons of each material were untreated (Control), treated with the ozone under standard methods (∼14 min. at ∼20 ppm O3), or treated with an enhanced ozone run (∼30 min. at ∼40 ppm O3). *Denotes a p-value where p < 0.01 when compared to control recoveries.

**Table 1.** Log Reductions of MS2 Bacteriophage from Coupons using the standard and enhanced treatments.

|  |  |  |
| --- | --- | --- |
| Material | Standard Treatment<br>Log-Reduction (S.D.) | Enhanced Treatment<br>Log-Reduction (S.D.) |
| 3M House<br>Dust Mask | 2.50 (0.194) | 2.38 (0.04) |
| N95 | 0.75 (0.383) | 3.38 (0.15) |
| Valve | 1.05 (0.283) | 1.43 (0.11) |
| Elastic Band | 3.95 (0.329) | 4.00 (NA) |

## Discussion

Various methods have been proposed for the decontamination of N95 masks for safe re-use by medical providers during the COVID-19 pandemic. Previous research has proposed the use of ultraviolet germicidal irradiation [8-10], ethylene oxide [8, 10]; vaporized hydrogen peroxide [8, 10-12]; microwave oven use [8, 9, 12]; bleach [13, 14], heat treatment [9, 10, 14], and liquid hydrogen peroxide [14].

In this study, we examined the efficacy of two different ozone treatments for the decontamination of household dust masks and N95 mask materials. Our results indicate that ozone treatment may be an effective method for decontamination of protective masks and respirators. Log-reductions of the MS2 phage were improved by lengthening the time and concentration of the ozone treatment consistent with other university testing. Using the enhanced cycle in the cabinet, the log-reductions of the MS2 phage from the N95 mask filter material and elastic were sufficient to meet the current guidelines for the decontamination of single-user masks and respirators for re-use, provided by the FDA (> 3 log10 reduction of non-enveloped viruses). Sufficient log reductions were not achieved to decontaminate the household dust mask or N95 exhalation valve to meet the FDA guidelines. The FDA guidance specifies that only approved N95 masks, without an exhalation valve, should be considered for reuse by healthcare workers.

Our data suggests that ozone treatment cabinets are a viable method for the decontamination and reuse of N95 respirators. Development of standardized methods for ozone sterilization has the potential to drastically increase the PPE capacity of the country allowing for increased access to highly effective FFRs and decreased cost burden to the user or organization.

## Limitations

While these results suggest ozone decontamination may be a viable method for PPE sterilization it should be noted this research is not comprehensive to the full range of commercially available FFRs. Further research is needed to evaluate the efficacy of these methods for different FFR filtration materials as well as additional PPE of interest (i.e., lab coats). Furthermore, due to the inability to obtain multiple masks at the time of this research, we were unable to determine if masks will preserve their integrity indefinitely or if ozone treatment will begin to degrade the material after repeated usage. However, we are aware of other research groups pursuing the degradation of FFRs with ozone decontamination. The limitation of mask availability also determined what sections of the masks we would focus on in this study. N95 masks are comprised of a variety of materials with different levels of porosity, making inoculation of MS2 variable between materials. The N95 exhalation valve is made of polypropylene & polyisoprene, relatively non-porous materials, which affected the drying efficiency of the inoculum on the material. However, it should be noted that masks with exhaust valves are not appropriate for reducing the spread of SARS-CoV-2 from the individual wearing the mask.

## Conclusion

The COVID-19 pandemic has dramatically reduced the availability of PPE. During this global emergency, decontamination and reuse of FFRs may be necessary when access to PPE is limited. We have demonstrated that ozone sterilization is an effective method for the decontamination of N95 mask materials. Decontamination of N95 mask materials with ozone sterilization cabinets resulted in > 3-log10 reduction in MS2, a conservative surrogate virus for SARS-CoV-2. Further research is needed to ensure that the integrity and performance of FFRs is maintained, even after decontamination with ozone.

## Data Availability

All data is available if requested.

